# Outbreak of sexually transmitted *S. sonnei bla*_CTX-M-15_ in England: an epidemiological and genomic investigation

**DOI:** 10.1101/2024.10.14.24314996

**Authors:** Hannah Charles, David R. Greig, Craig Swift, Israel Olonade, Ian Simms, Katy Sinka, Kate S Baker, Gauri Godbole, Claire Jenkins

**Affiliations:** Blood Safety, Hepatitis, STI and HIV division, UK Health Security Agency, London, UK; Gastrointestinal Bacteria Reference Unit, UK Health Security Agency, London, UK; NIHR Health Protection Research Unit in Gastrointestinal Infections, University of Liverpool, Liverpool, UK; Department of Genetics, School of Biological Sciences, University of Cambridge, Cambridge, United Kingdom

## Abstract

The diarrhoeal disease, shigellosis, can be sustained as a sexually transmissible enteric illness among gay, bisexual, and other men who have sex with men (GBMSM). Multiple extensively drug-resistant strains of *Shigella* have been detected through genomic surveillance, which have typically been associated with plasmids carrying the gene variant *bla*_CTX-M-27_. We report an increase in likely sexually transmissible cases of *Shigella* carrying *bla*_CTX-M-15,_ which was previously associated with travel. In 2023, there were 117 cases belonging to the single 10-SNP single linkage cluster, t10.1814. While this cluster had been present in England since August 2019, genetic analyses revealed that *bla*_CTX-M-15_ entered the lineage on a novel resistance plasmid coincident with the first case of the outbreak. This highlights the shifting antimicrobial resistance landscape of sexually transmissible *Shigella* and the parallel emergence of resistance determinants against third generation cephalosporins in sexual transmission networks suggests high levels of antimicrobial selection pressure among GBMSM.

## Introduction

Shigellosis is a gastrointestinal infection caused by one of four bacterial species, *Shigella sonnei, S. flexneri, S. boydii* or *S. dysenteriae*. The most common symptoms are bloody diarrhoea, abdominal pain and cramps, and fever, nausea and/or vomiting (1). *Shigella* species are anthroponotic, and transmitted via faecal-oral contact (1), either on hands or on objects which have been in contact with human faeces, including through sexual contact (2). Infection can also occur via contaminated food and water (3, 4), and travel to endemic countries (such as those in sub-Saharan Africa, Southern Asia and Latin America) (5). Community outbreaks have been associated with childcare settings, schools, residential institutions, and restaurants (6-8). People at highest risk of infection include those attending childcare settings, travellers to endemic countries and gay, bisexual and other men who have sex with men (GBMSM).

The implementation of whole genome sequencing (WGS) for public health surveillance of bacterial pathogens has facilitated monitoring of the emergence and transmission of epidemic strains of *Shigella* spp., as well as antimicrobial resistance (AMR) on a global scale. AMR *S. sonnei* were first described over 60 years ago (9, 10), and multi-drug resistant (MDR) strains resistant to aminoglycosides, sulphonamides, trimethoprim and/or chloramphenicol are endemic in the human population on every continent (11). Resistance to fluroquinolones has recently emerged, including from regions where antimicrobial use is unregulated (12-14). The increasing incidence of MDR and extensively-drug resistant (XDR) shigellosis in high-prevalence regions where surveillance is limited can be monitored by sequencing strains of *S. flexneri* and *S. sonnei* isolated from returning travellers and analysing the genome derived AMR profiles.

Since 2010, surveillance systems maintained by the UK Health Security Agency (UKHSA) have identified a series of epidemics of MDR *S. flexneri* serotypes 3a, 2a and 1b and *S. sonnei* among GBMSM circulating nationally and internationally (11, 15-17). Previous studies demonstrated that acquisition of a plasmid encoding resistance to macrolides corresponded with the emergence of the sequential epidemics of *S. flexneri* 3a, 2a and *S. sonnei* between 2010 and 2015 (18, 19). The subsequent global increase in notification of *S. sonnei* among GBMSM was facilitated by strains belonging to global lineage 3.6.1.1.2 exhibiting resistance to both macrolides and fluroquinolones (15). During the COVID-19 pandemic, in the UK there was a rapid decrease in notifications of *S. sonnei*. However, following the relaxation of social distancing and travel restrictions, notification quickly returned to pre-pandemic levels (20), and we observed an increase in the proportion of XDR *S. sonnei* harbouring *bla*_CTX-M-27_, conferring resistance to third generation cephalosporins (21). Outbreaks of XDR *S. sonnei* containing *bla*_CTX-M-27_ *and S. flexneri* containing *bla*_CTX-M-27_, primarily circulating within GBMSM sexual networks, have been described previously (17, 22), however, these outbreaks were relatively localised and short-lived. In contrast, the epidemic of sexually transmitted XDR *S. sonnei* was recognised in September 2021 (designated t10.377, contained within global lineage 3.6.1.1.2), continued into 2022 and been reported internationally (21).

Following the publication of a French study reporting an increase in the proportion of *Shigella* spp. isolates simultaneously resistant to ciprofloxacin, third generation cephalosporins and azithromycin (23), we reviewed genome-derived AMR profiles of the *S. sonnei* in the UKHSA archive isolated between 2016 and 2023. We identified an increasing trend of XDR strains of *S. sonnei* and found that XDR *S. sonnei* isolated from MSM almost exclusively had the *bla*_CTX-M-27_ variant, whereas XDR *S. sonne*i isolated from travellers returning from high-risk regions almost exclusively had the *bla*_CTX-M-15_ variant (24). In 2023, we detected a notable increase of XDR *S. sonnei* in England that contained *bla*_CTX-M-15_. The aim of this study was to use a combination of epidemiological data and short-read and long-read genomic sequencing data to investigate the outbreak to elucidate the emergence and transmission patterns of the outbreak strain and the acquisition of *bla*_CTX-M-15_.

## Methods

### Routine laboratory and epidemiological surveillance

*Shigella* spp. isolates from hospital and community cases with gastrointestinal symptoms are referred to the Gastrointestinal Bacterial Reference Unit (GBRU) at the UKHSA for confirmation and typing. Since 01 September 2015, WGS has been conducted for all *Shigella* spp. isolates submitted to the GBRU, where the serotype and AMR profile are derived *in silico* from the genome, using methods previously described (25). *S. sonnei* isolates submitted to the GBRU from 01 January 2016 to 31 December 2023 were included in this study. Given the lack of sexual orientation information available in this dataset, in line with previous work, we used a proxy indicator of cases that may be attributed to sexual transmission among GBMSM, defined as cases among adult males without a history of travel or where travel history was unknown (herein presumptive MSM) (26).

We analysed the sequencing data for genomic markers of resistance to azithromycin (defined here as the presence of *ermB* and/or *mphA*), ciprofloxacin (defined here as the presence of mutations in *gyrA, parC and/or qnr*), and third generation cephalosporins (defined here by the presence of *bla*_CTX-M-15_ or *bla*_CTX-M-27_). We defined extensively-drug resistant isolates as those containing genomic markers of resistance to azithromycin, ciprofloxacin and third generation cephalosporins.

Single-nucleotide polymorphism (SNP) typing was carried out on *S. sonnei* isolates. Single-linkage hierarchical clustering was applied at seven descending thresholds of SNP distances (Δ250, Δ100, Δ50, Δ25, Δ10, Δ5, Δ0) as previously described (26). This clustering results in a discrete seven-digit code where each number represents the cluster membership at each descending SNP distance threshold. For the purpose of *Shigella* spp. surveillance, isolates that cluster at the 10 SNP threshold are designated t10.X. Sequencing data were de-duplicated in-line with routine genomic surveillance of *Shigella* spp. at UKHSA. Differences in proportions were tested using two-proportion Z-tests, with the significance threshold set at 0.05.

### Phylogenetic tree construction

The WGS data from routine laboratory surveillance at the UKHSA were utilised to conduct a phylogenetic tree of *S. sonnei* isolates harbouring *bla*_CTX-M_ variants. A soft-core genome alignment of Clonal Complex 152 (CC152) t25:1, where a given variant position that belonged to a minimum of 80% of strains in the alignment, totalling 1,325 samples, was produced from SnapperDB v0.2.8 (27). Recombinant sequences were previously masked by Gubbins v3.2 (28) from a whole genome alignment derived from SnapperDB v0.2.8 (27) on the same dataset. The alignment (totalling 2,142,354 bases in size) was used as the input for IQ-TREE v2.0.4 (29) to generate a phylogenetic tree. We then repeated the methodology to produce subtrees of each of the clusters containing genomes harbouring *bla*_CTX-M_ variants.

### Nanopore sequencing and *de novo* assembly

In addition to routine Illumina-based sequencing, long-read sequencing using Oxford Nanopore Technologies (ONT) was used to generate complete assemblies of selected *bla*_CTX-M_ variants samples to determine the genetic context for antimicrobial resistance determinants. Genomic DNA was extracted and sequenced using the MinION, data processed, read trimming and assembly as described previously (30).

*De novo* assembly was performed using Flye v2.9.2 (31). Correction of the assemblies was performed in a two-step process; firstly Medaka (v1.0.3) (https://github.com/nanoporetech/medaka) with a *Shigella*-specific medaka-trained model. Secondly, using Polypolish v0.5.0 (32) using the equivalent Illumina FASTQs for each assembly.

Finally, as all contigs were circular and closed, they were reoriented to start at the *dnaA* gene (NC_000913) from *E. coli* K12, using the --fixstart parameter in Circlator v1.5.5 (33).

### AMR gene detection and plasmid typing

The plasmid replicon for each non-chromosomal contig within the final assembly of each sample was detected using PlasmidFinder v.2.1 (34) with the *Enterobacteriaceae* database and the following parameters set: minimum identity=90% and minimum coverage = 90%. The mobile genetic elements (MGEs) harbouring AMR determinants were annotated using PGAP build 2022-12-13 (35) Finally, gene-level alignments were generated using Clinker v.0.0.27 (36).

### Data deposition

All FASTQ and assemblies were submitted to the National Centre for Biotechnology Information (NCBI) under NCBI BioProject: PRJNA315192 and can be found in Supplementary Table 1.

### Ethical statement

This analysis was undertaken for health protection purposes under permissions granted to UKHSA to collect and process confidential patient data under Regulation 3 of The Health Service (Control of Patient Information) Regulations 2020 and Section 251 of the National Health Service Act 2006.

## Results

### Descriptive epidemiology

Overall, diagnoses of *S. sonnei* increased between 2016 and 2018, before declining slightly in 2019 and then further declining markedly in 2020 and 2021, likely due to reduced access to healthcare services and testing, and social distancing and travel restrictions, during the COVID-19 pandemic (Figure 1). In 2022 and 2023, there was a substantial increase in *S. sonnei* diagnosis notifications, such that the number in 2023 exceeded pre-pandemic levels. The trends of *S. sonnei* among presumptive MSM mirror those among all persons. However, the rate of increase was larger among presumptive MSM, leading to an increase in the proportion of all *S. sonnei* diagnoses that were seen among presumptive MSM, from 26% in 2016 to 46% in 2023. The increase between 2022 and 2023 was also higher among presumptive MSM compared to all persons (82% increase compared to a 50% increase).

**Figure 1.**
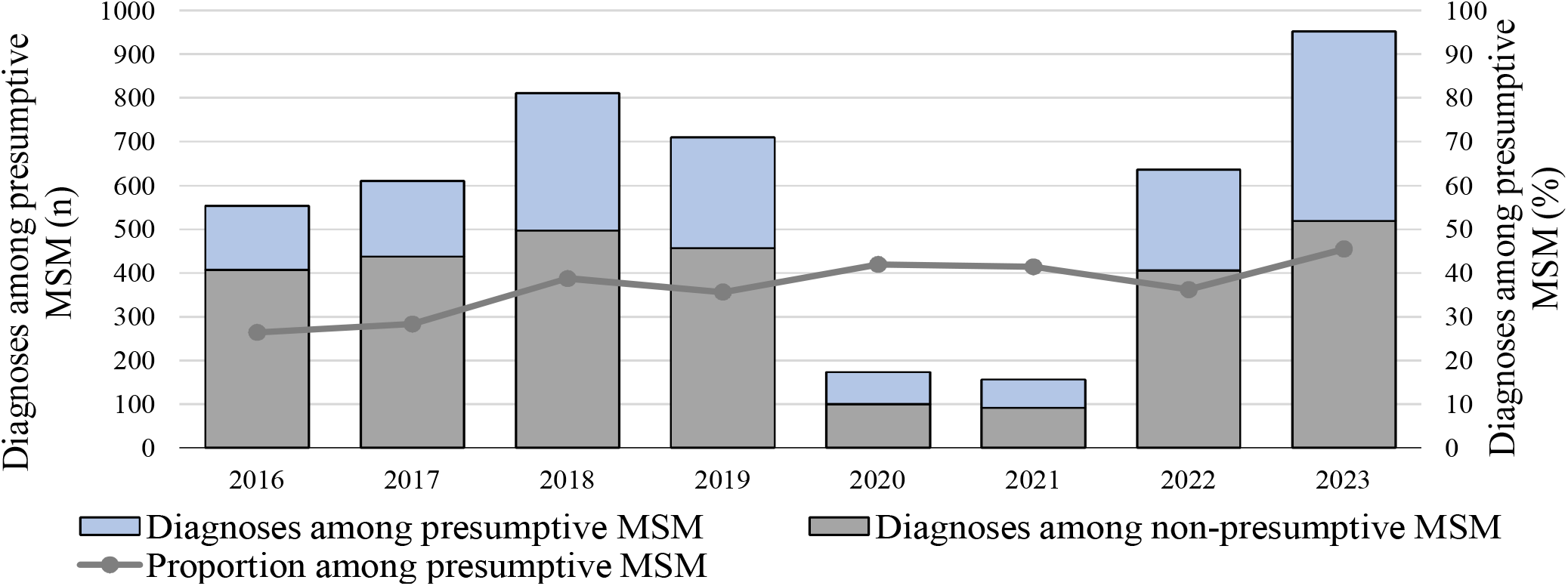
*S. sonnei* diagnoses by presumptive MSM classification, 2016 to 2023, England.

As well as the increase in *S. sonnei* among presumptive MSM in 2023, there was a corresponding increase in the number and proportion of *S. sonnei* harbouring *bla*_CTX-M-15_ in this population group. Between 2016 and 2022, an average of 10% of *S. sonnei* isolates contained *bla*_CTX-M-15_, increasing to 33% in 2023. This increase in the proportion of *S. sonnei* isolates with *bla*_CTX-M-15_ in 2023 corresponded with a decrease in *S. sonnei* isolates with *bla*_CTX-M-27_ in this population group (Figure 2).

**Figure 2.**
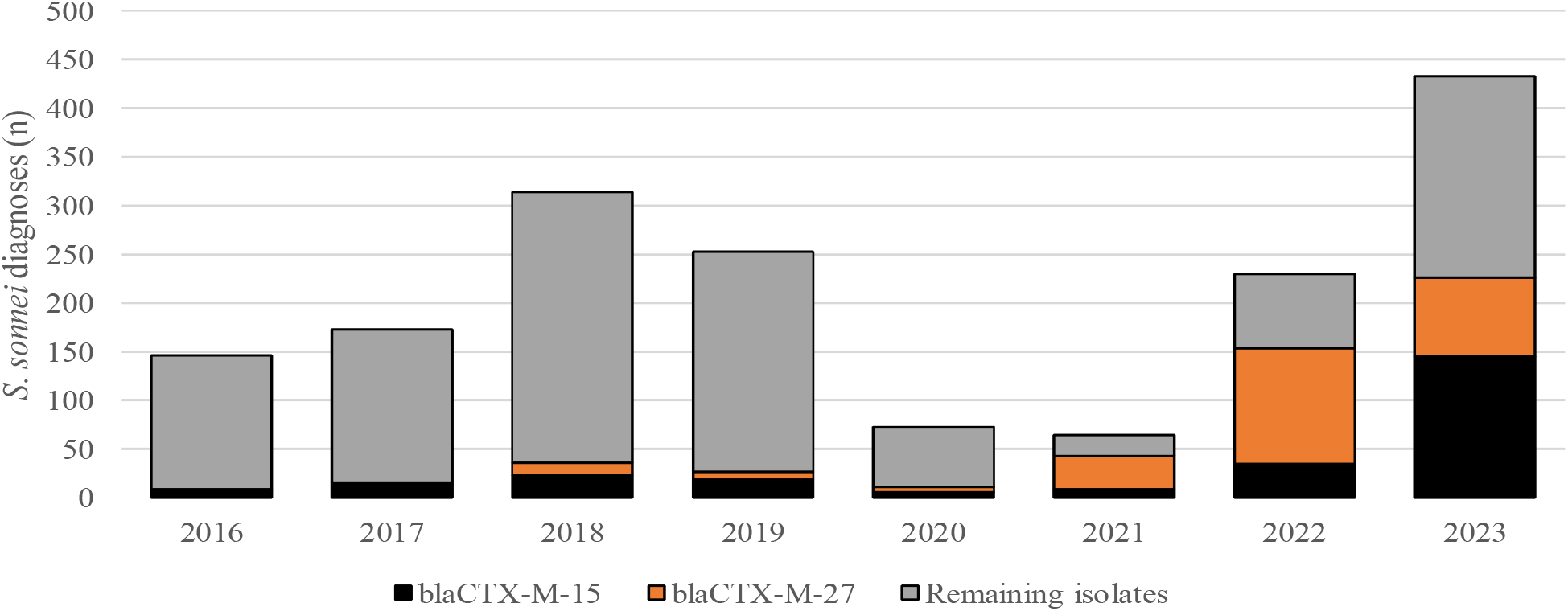
*S. sonnei* isolates among presumptive MSM by presence of *bla*_CTX-M-15_ or *bla*_CTX-M-27_, 2016 to 2023, England.

Prior to 2023, *S. sonnei* isolates with *bla*_CTX-M-15_ were identified at a much lower frequency among presumptive MSM compared to non-presumptive MSM (females, children and men reporting recent travel). Between 2016 and 2022, the proportion of *S. sonnei* with *bla*_CTX-M-15_ among presumptive MSM remained stable at an average of 17%, increasing to 38% in 2023 (Figure 3).

**Figure 3.**
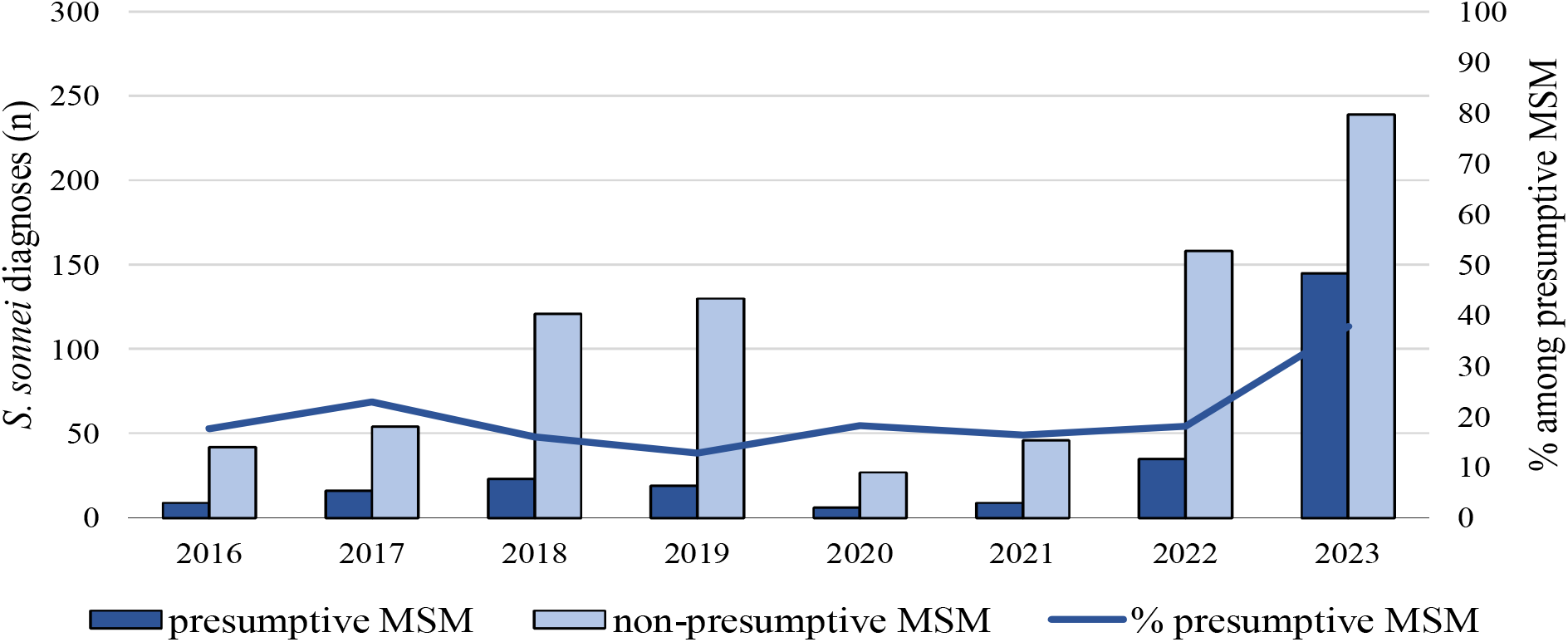
*S. sonnei* isolates with *bla*_CTX-M-15_ among presumptive MSM compared to non-presumptive MSM, 2016 to 2023, England.

### Phylogenetic analysis of *S*. *sonnei* harbouring *bla*_CTX-M-15_

Of the 262 *S. sonnei* isolates among presumptive MSM harbouring *bla*_CTX-M-15_ between 2016 and 2023, 84 (32%) fell within a single 10 SNP single linkage cluster (SLC) designated t10.1814 (1.1.1.1.1814.%) and belonging to global subtype 3.6.1.1 (37) (Figure 9). In addition, two other 10 SNP SLCs contained isolates harbouring *bla*_CTX-M-15_ were identified (1.1.29.49.1148.% and 1.1.29.49.2187.%), these two clusters fall within the same 25 SNP SLC (t25:49), a lineage which is associated with travel to Pakistan (82% of cases reporting travel to Pakistan) and Tunisia (94% of cases reporting travel to Tunisia) respectively, but distinct from the 25 SNP SCL containing t10.1814 (t25:1). The remaining *S. sonnei* isolates among presumptive MSM harbouring *bla*_CTX-M-15_ were phylogenetically sporadic and did not form large clusters (Figure 6). Therefore, our phylogenetic analysis focuses on the t10.1814 cluster to explore the increase in *S. sonnei* harbouring *bla*_CTX-M-15_ among presumptive MSM (Figure 9).

At the end of 2023, the t10.1814 cluster contained 124 isolates in total. The first three cases within this cluster were diagnosed in August and October 2019. None of these isolates contained *bla*_CTX-M-15_, however two out of the three isolates contained *bla*_CTX-M-27_ (Figure 4, Figure 6, Figure 9). There was no reported activity within the t10.1814 cluster until March 2022, when there were a further four cases between March and December 2022. There was a substantial increase in cases in 2023 such that almost all isolates in this cluster had specimen dates in 2023 (117 out of 124; 94%). Of the isolates in 2023, 92% harboured *bla*_CTX-M-15_ (108 out of 117) (Figure 4).

**Figure 4.**
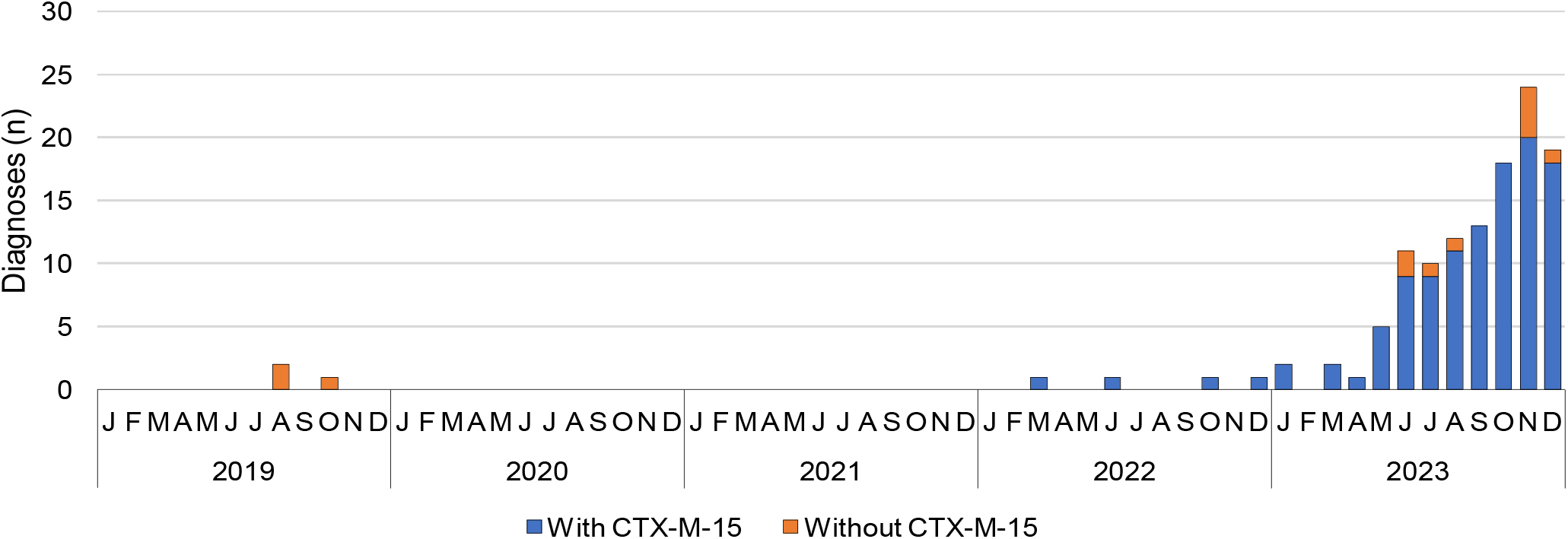
Diagnoses of *S. sonnei* in cluster t10.1814 by *bla*_CTX-M-15_ status, 2019 to 2023, England.

A high proportion of cases within this cluster were adult males (113 out of 124; 91%) who had not reported travel outside the UK or for whom travel history was unknown (104 out of 124; 84%).

Twenty cases (16%) reported travel, the majority of which was to European countries (Table 1).

**Table 1:** Characteristics of cases within t10.1814 cluster, 2016 to 2023, England.

|  |  | N=124<br>n (%) |
| --- | --- | --- |
| Gender | Male | 113 (91) |
|  | Female | 11 (9) |
| Age | Median [IQR] | 35 years [30-43] |
|  | Adult | 123 (99) |
|  | Child | 1 (1) |
| Travel | Yes | 20 (16) |
|  | No/Unknown | 104 (84) |
| Travel destination (of those reporting travel) | Belgium | 1 (5) |
|  | Brazil | 1 (5) |
|  | Europe (country unknown) | 1 (5) |
|  | Germany | 1 (5) |
|  | India | 1 (5) |
|  | Netherlands | 1 (5) |
|  | North America | 1 (5) |
|  | Spain | 8 (40) |
|  | Thailand | 1 (5) |
|  | United states | 1 (5) |
|  | Unknown | 3 (15) |
| Presumptive MSM | Yes | 93 (75) |
|  | No | 31 (25) |

The increase in t10.1814 occurred in parallel to a decline in cases within the 1.1.1.1.377.% cluster (designated t10.377); a cluster that had been dominant since 2017, declined substantially during the first few months of the COVID-19 pandemic but re-emerged in September 2021 with third generation cephalosporin resistance via *bla*_CTX-M-27_ (21) (Figure 5).

**Figure 5.**
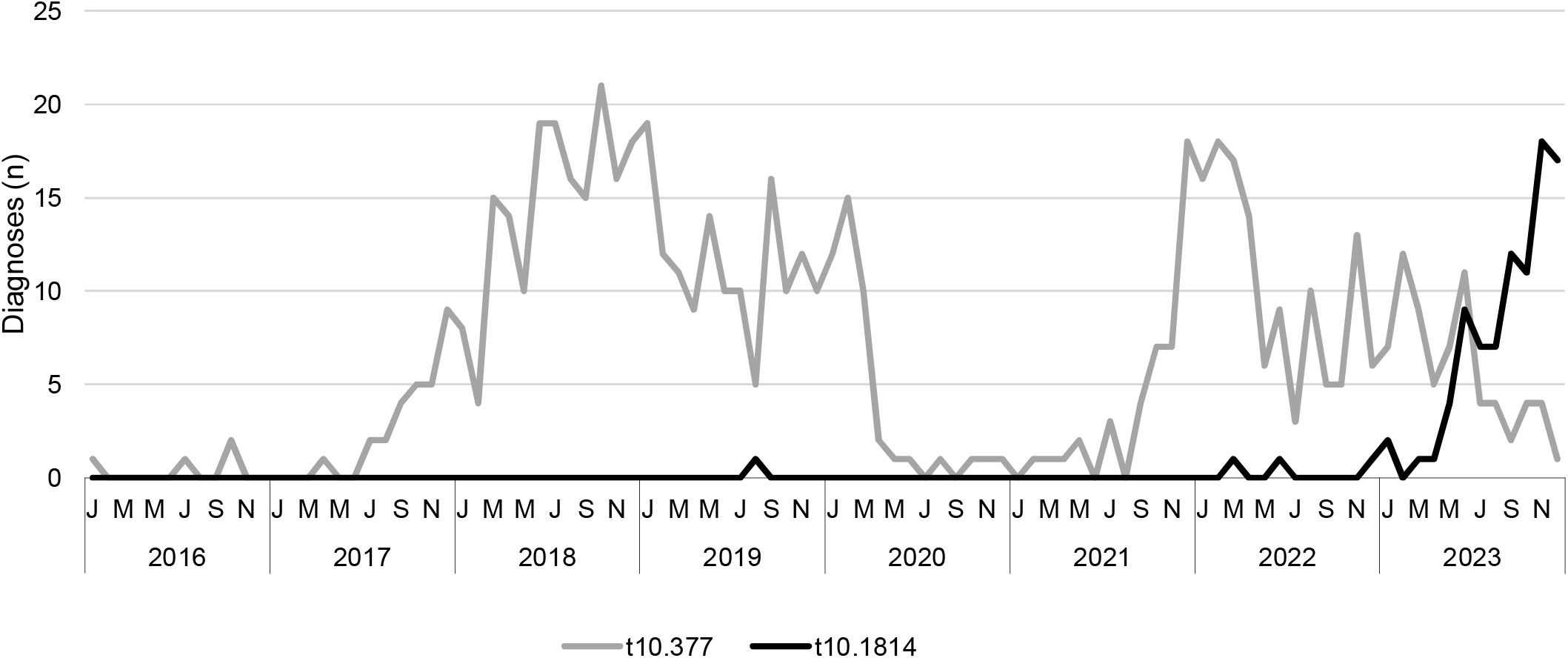
Diagnoses of *S. sonnei* in t10.377 cluster compared to t10.1814 cluster among presumptive MSM, 2016 to 2023, England.

Almost all isolates in the t10.1814 cluster harboured *bla*_CTX-M-15_ (112/124; 90%), whilst only two isolates harboured *bla*_CTX-M-27_; there were no isolates expressing both *bla*_CTX-M-15_ and *bla*_CTX-M-27_. Resistance to ciprofloxacin and azithromycin were similarly very high – 94% of isolates (116/124) exhibited either mutations in *gyrA* and/or *parC* and/or the plasmid mediated *qnr* gene variant and 88% (109/124) had genomic markers for azithromycin resistance (*ermB* and/or *mphA*) (Table 2). 107 out of 124 (86%) isolates had both *bla*_CTX-M-15_ and markers of azithromycin resistance.

**Table 2:**
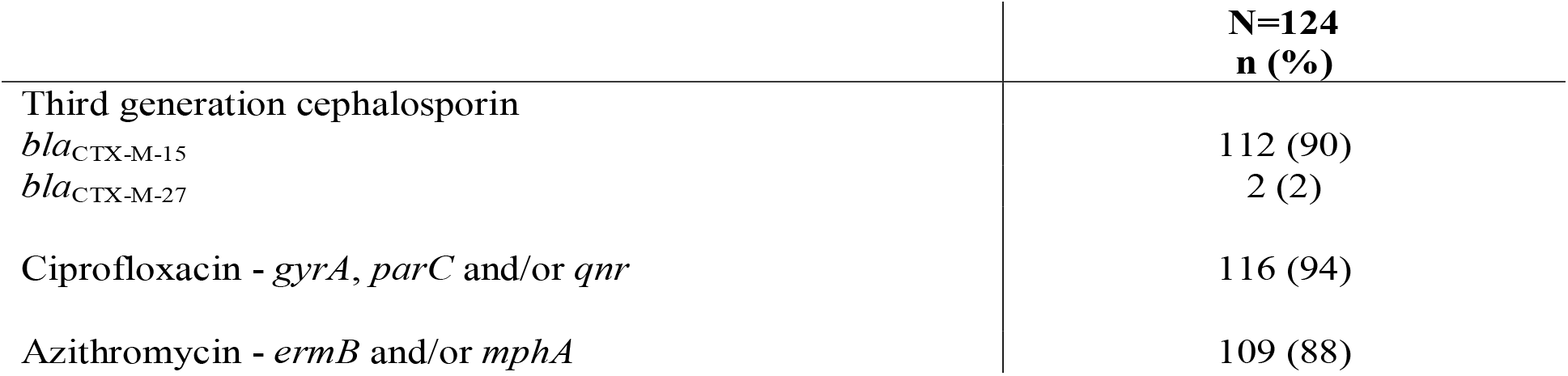
Antimicrobial resistance profile of cases within the t10.1814 cluster, 2016 to 2023, England.

|  | **N=124**<br>**n (%)** |
| --- | --- |
| Third generation cephalosporin |  |
| *bla*<sub>CTX-M-15</sub> | 112 (90) |
| *bla*<sub>CTX-M-27</sub> | 2 (2) |
| Ciprofloxacin - *gyrA*, *parC* and/or *qnr* | 116 (94) |
| Azithromycin - *ermB* and/or *mphA* | 109 (88) |

Phylogenetic analysis revealed that t10.1814 fell within the wider t25:1 cluster which also includes t10.377. Although located within the same t25 single linkage cluster, the t10.1814 cluster harbouring *bla*_CTX-M-15_, described here, was located on a separate branch and did not evolve from the t10.377 cluster harbouring *bla*_CTX-M-27_ (Figure 6). The progenitor strains of t10.1814 cluster harbouring *bla*_CTX-M-15_ contained *bla*_CTX-M-27_.

**Figure 6.**
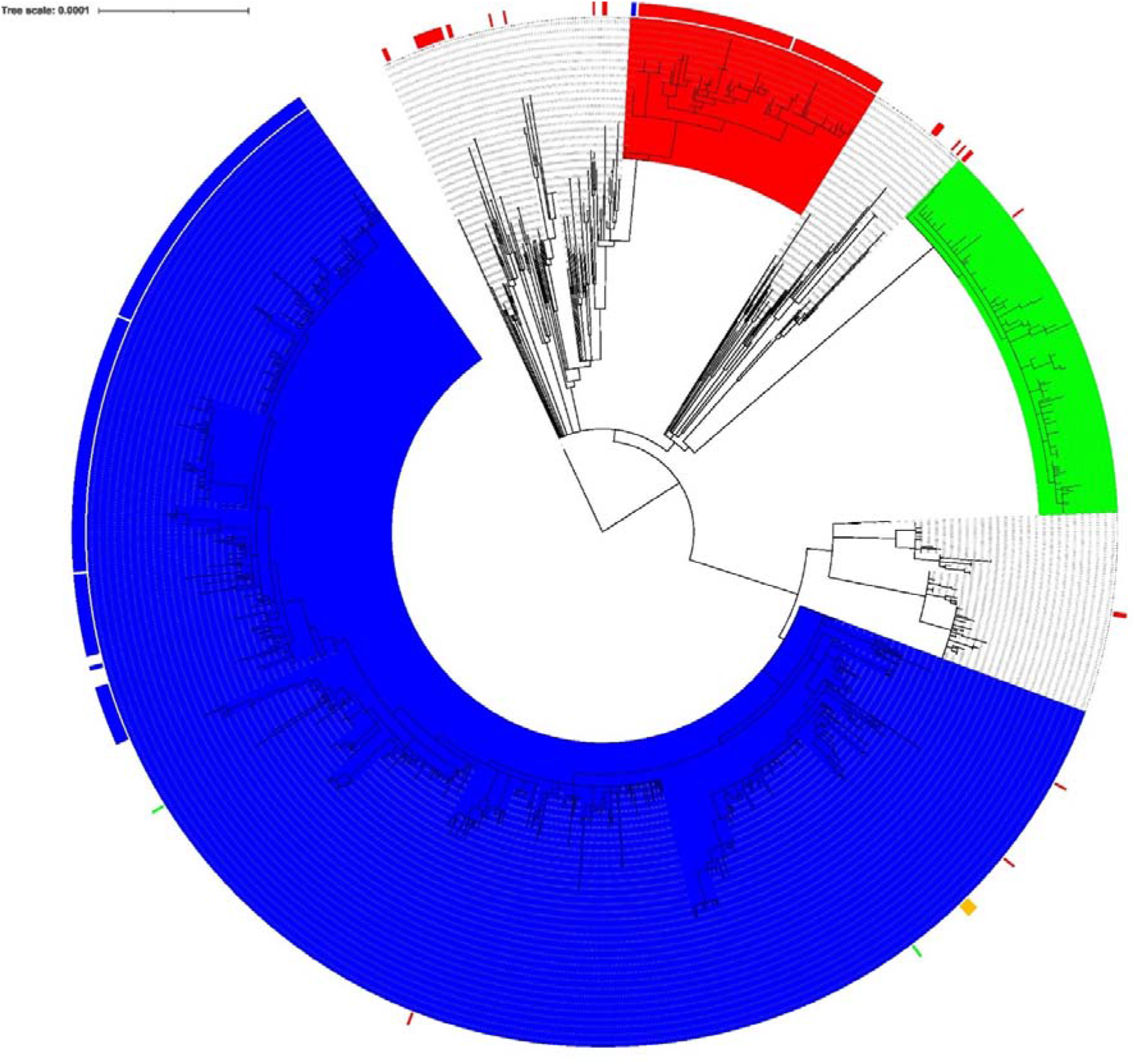
Maximum-likelihood phylogenetic tree of CC152 t25:1 (n=1,325) showing t10.377 cluster in blue, t10.2218 in green and t10.1814 in red. The outer ring indicates presence of *bla*_CTX-M_ gene; where red is *bla*_CTX-M-15_, blue is *bla*_CTX-M-27_, orange is *bla*_CTX-M-134_ and green is *bla*_CTX-M-55._

### Analysis of t10.1814 IncFII plasmids and comparison to t10.377 IncFII plasmids

Plasmids within t10.1814 harbouring *bla*_CTX-M-15_ were all determined to be of the IncFII replicon type and ranged from 77.6kbp to 149.0kbp in size (Figure 7). Plasmids from progenitor strains within t10.1814 harbouring *bla*_CTX-M-27_ were larger (average 148kbp) than those harbouring *bla*_CTX-M-15_ (78.4kbp). Despite the difference in size, almost all gene content of the ∼74kbp plasmids were also found within the larger ∼148kbp plasmids.

**Figure 7.**
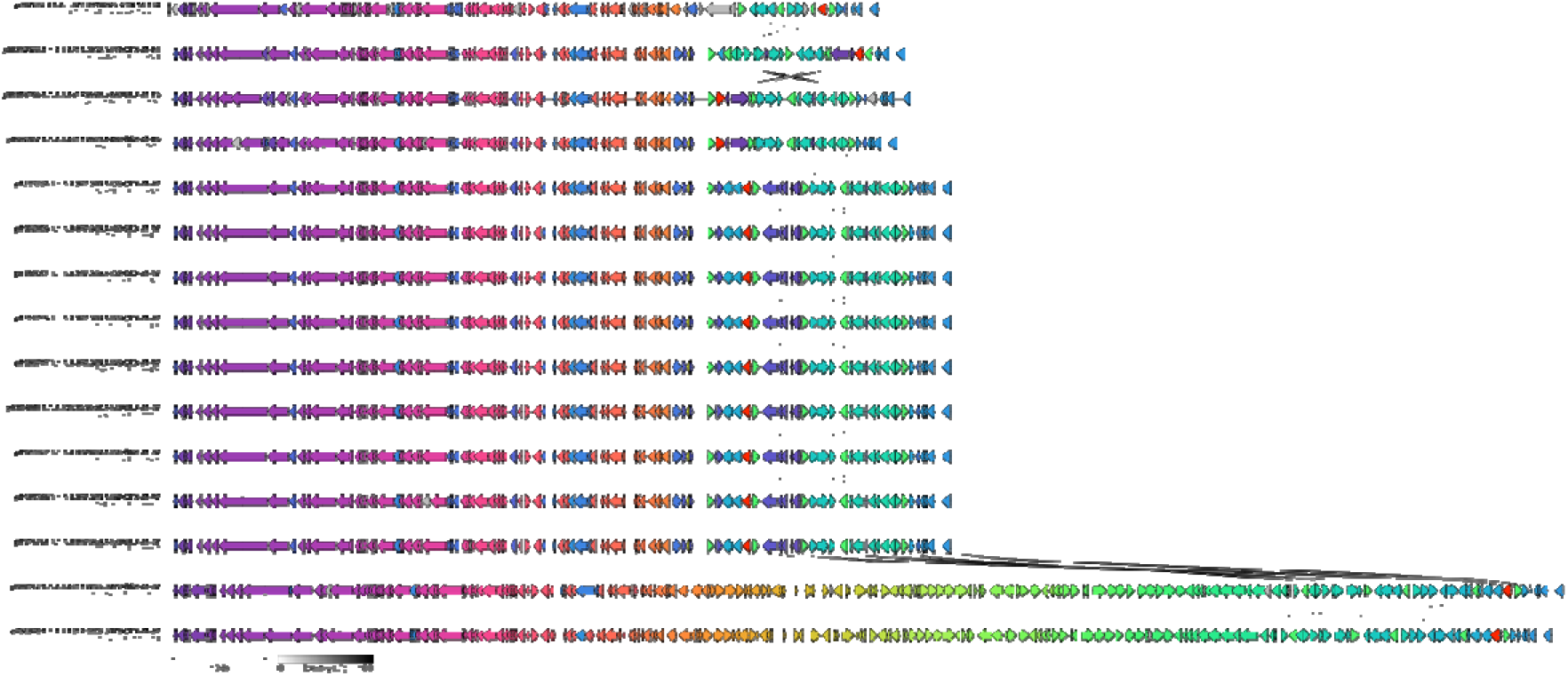
Alignment of IncFII plasmids in samples selected for Nanopore sequencing. Gene coloured red indicates the *bla*_CTX-M_ variant.

When comparing t10.1814 plasmids to IncFII plasmids from t10.377, it was noted that the plasmid structure is the same, except for small alterations to the variable region, including the *bla*_CTX-M-15_ and *bla*_CTX-M-27_ integrons, respectively (Figure 7).

Following the analysis of the long read sequencing data, we identified one isolate of *S. sonnei* harbouring *bla*_CTX-M-15_ in the same clade as t10.1814, but in a different t10 SLC cluster (t10.2404) where *bla*_CTX-M-15_ was located on a 7.6 kbp cassette/element integrated on the chromosome. The integration site appears to a prophage remnant (a region where multiple prophages have integrated and vacated leaving behind several genes) near a tRNA-Phe gene (Figure 8). The IncFII plasmid (known to encode *bla*_CTX-M-15_ or *bla*_CTX-M-27_) in other isolates, had been lost in this sample.

**Figure 8.**
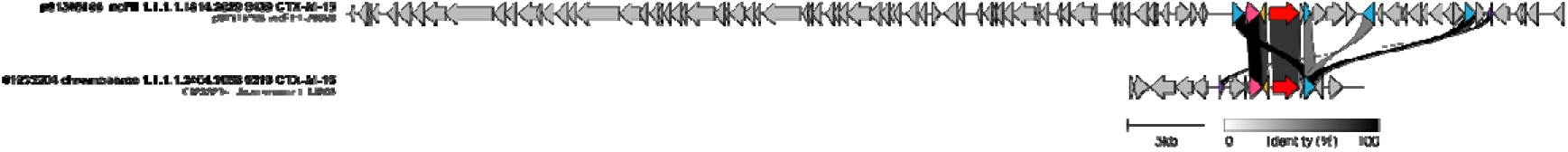
Alignment of exemplar IncFII plasmid from a strain within t10.1814 and strain 01233204 (SRR29176725) showing the cassette containing *bla*_CTX-M-15_ (gene highlighted red) has moved to the chromosome.

**Figure 9.**
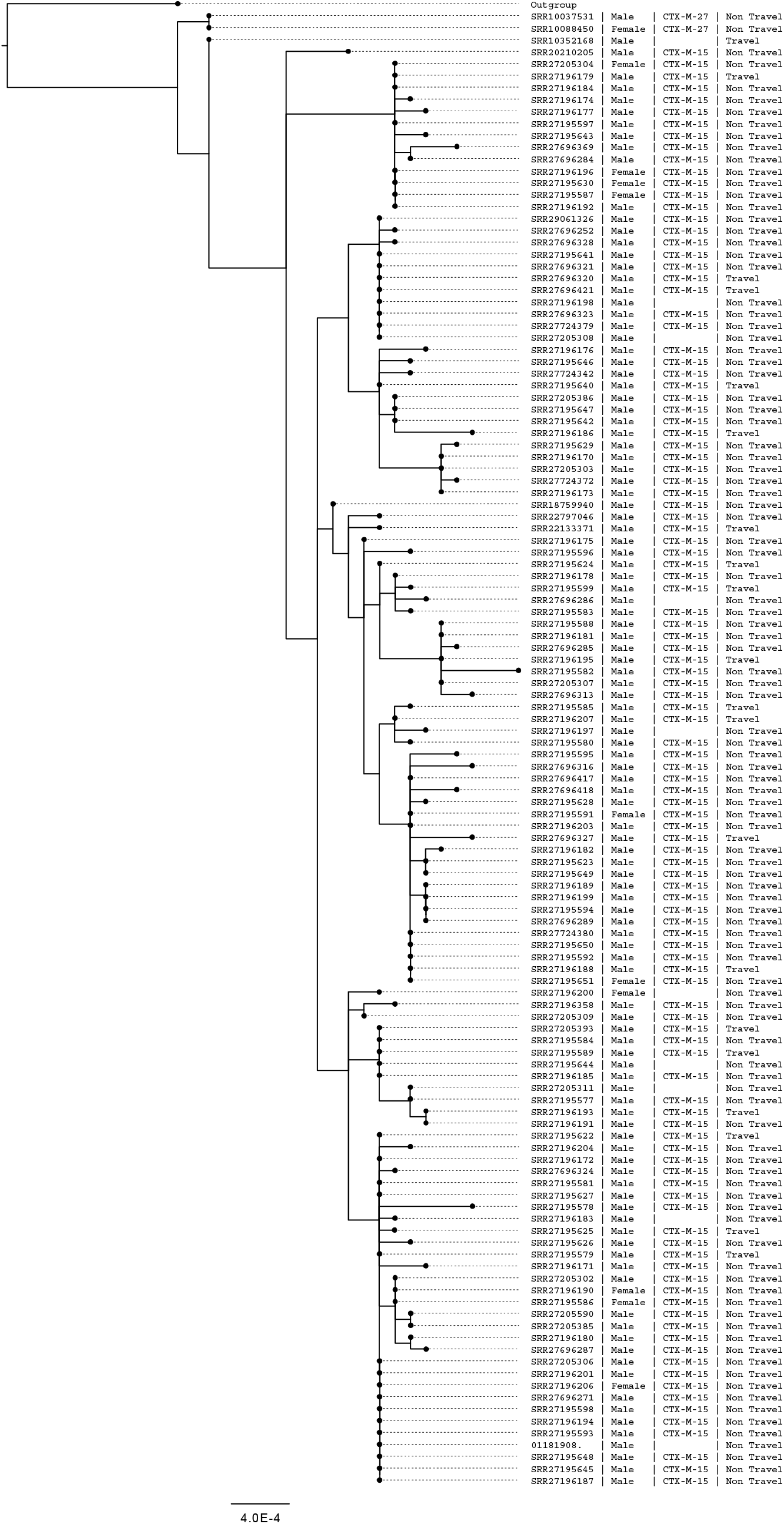
Maximum-likelihood phylogenetic tree of CC152 t10.1814 (n=125). Detailing sample ID as SRR accession, gender, presence of *bla*_CTX-M_ variants and association with travel.

## Discussion

Overall, with the exception of 2020 and 2021 when notifications were impacted by social distancing and travel restrictions, we observed a steady increase of *S. sonnei* diagnoses in England between 2016 and 2023. Over the last decade, diagnostic methods for the detection of gastrointestinal pathogens have improved with the widespread implementation of commercial polymerase chain reaction (PCR) assays. PCR is more sensitive than culture for the detection *Shigella* spp. (38, 39), and this move towards molecular methods followed by culture will have increased case ascertainment. Furthermore, the increase may be associated with increased travel to high-risk regions, although this is difficult to confirm as travel history is poorly captured by the current surveillance system. We also observed a steady increase in notifications of *S. sonnei* among presumptive MSM. Whilst this may reflect a true increase in sexual transmission, it may also be influenced by the factors described above. The increase in reported diagnoses may also be a consequence of the publication of briefing notes and other outbreak-related communications during the period of this analysis by UKHSA.

There are numerous factors facilitating the emergence, transmission, and persistence of epidemic strains circulating within GBMSM sexual networks, involving pathogen characteristics, host behaviours, and/or environmental pressures. We have previously hypothesised that the sequential waves of shigellosis among GBMSM in the UK have been facilitated by acquisition of AMR to an increasing number of classes of antimicrobials (40). With respect to *S. sonnei*, the epidemic strains were initially resistant to macrolides, then to both azithromycin and ciprofloxacin, and most recently to macrolides, fluroquinolones and third generation cephalosporins (15, 21). However, acquisition of AMR alone does not explain the emergence and persistence of all shigellosis epidemics among GBMSM. For example, in this study, we showed that the previous epidemic *S. sonnei* strain (t10.377) was replaced by another strain of *S. sonnei* (t10.1814) and with the same genotypic AMR profile, and the re-emergent strain of *S. flexneri* serogroup 3a in 2019 was more susceptible to antibiotics than the strain that caused the original *S. flexneri* 3a epidemic (41). During previous epidemics, there are examples of the emergence and persistence of strains outcompeting other strains exhibiting the same AMR profiles and has been suggested that other factors might be at play, such as transient host immunity to circulating serotypes may provide emergent serotypes with a competitive advantage. Host immunity seems an unlikely explanation for the strain replacement event described here as both strains were *S. sonnei*.

Overall, the characteristics of cases in the t10.1814 cluster were similar to those in the t10.377 cluster in terms of the proportion of male cases and age distribution. There is some regional variation, with cases in the t10.1814 cluster being more dispersed across regions of England, and the t10.377 cluster being more concentrated in London (Supplementary Table 2). The difference in travel history between cases in these clusters could be an artefact of the large proportion of missing data on recent travel history.

Phylogenetic analyses showed clustering of *bla*_CTX-M_ variants within the *Shigella* population structure, consistent with horizontal acquisition and subsequent vertical transmission. Non-GBMSM clades associated with *bla*_CTX-M-15_ comprised cases reporting travel to high-risk regions, highlighting the possibility that this resistance determinant was brought in through travel, as speculated for with *Shigella* in other regions (42, 43). There was one GBMSM *bla*_CTX-M-15_ clade comprising isolates that fell within the same 10 SNP single linkages cluster, and although the *bla*_CTX-M-27_ decrease coincided with the *bla*_CTX-M-15_ increase, there was no evidence that *bla*_CTX-M-15_ had emerged from the clade harbouring *bla*_CTX-M-27_ The acquisition of the *bla*_CTX-M-15_ therefore appears to be an independent evolutionary event on a different branch of the phylogeny.

Long read sequencing analysis revealed that, like the *bla*_CTX-M-27_ determinant in the t10.377 cluster, *bla*_CTX-M-15_ in the current epidemic cluster, t10.1814, was located on an IncFII plasmid. Despite encoding different *bla*_CTX-M_ variants, the plasmid encoding *bla*_CTX-M-15_ exhibited high levels of similarity to the plasmid encoding *bla*_CTX-M-27_. These data show that similar IncFII plasmids persist and remain stable in the strains of *S. sonnei* circulating among GBMSM, despite acquisition of different AMR determinants. Given this apparent plasmid stability in this population, and our demonstration of the acquisition of *bla*_CTX-M-27_ and *bla*_CTX-M-15_ and subsequent clonal expansion, there remains the potential that other AMR determinants could be acquired onto this plasmid, and worsen the already concerning AMR picture of *S. sonnei*. In addition, we report an isolate in a separate clade (t10.2404) where the *bla*_CTX-M-15_ gene was located on the chromosome and the associated plasmid had been lost.

Social distancing and travel restrictions in 2020 and 2021 related to the COVID-19 pandemic had a greater impact on reducing notification of *S. sonnei* than *S. flexneri* (24). Previously, we considered that globalisation and increased travel may have a role in seeding sexually transmissible shigellosis. The acquisition of the *bla*_CTX-M-15_ determinant previously associated with travel-related cases of *S. sonnei*, on the GBMSM-associated IncFII pKSR-100-like plasmid may provide further evidence for this hypothesis. The reporting of *S. sonnei* harbouring *bla*_CTX-M-15_ among GBMSM in several other European countries further suggests the potential international distribution of this lineage (44, 45).

This study has some limitations. With a lack of information about sexual orientation, and incomplete travel histories, it is possible that adult male cases who travelled were categorised as presumptive MSM within this cluster if these travel histories were not known. Identifying as GBMSM and reporting recent travel are also not mutually exclusive, therefore there are limitations with the use of the presumptive MSM proxy definition. It is also not mandatory for primary diagnostic laboratories to send *S. sonnei* isolates to the GBRU, so the data available for this analysis represents a proportion of the total number of reported infections (approximately two thirds).

Despite these limitations, the introduction of WGS for typing gastrointestinal pathogens greatly improved surveillance of *S. sonnei* at UKHSA. Previously we relied on phenotypic methods that were highly specialised, labour intensive and difficult to standardise, such as phage typing and antibiotic susceptibility testing. Over the last decade, sequencing data has been utilised to construct the population structure of *S. sonnei* from UK residents and mapped clades associated with travel and those associated with sexual transmission among GBMSM. We have tracked the rise and fall of different clades circulating within GBMSM sexual networks and showed that acquisition of AMR and genetic factors contribute to emergence, transmission and persistence. However, notifications continue to rise, and the circulating strains are increasingly resistant to first- and second-line antimicrobials.

This work highlights the continued utility of genomic surveillance in detecting outbreaks of sexually transmissible shigellosis. Furthermore, through detailed analyses of the data we can understand the complex origins and transmission pathways for antimicrobial resistance in increasingly antimicrobial resistant strains. We now recurrently see conjugative plasmids carrying resistance against key antimicrobial classes mobilising among *Shigella* strains circulating in different transmission networks. This underlines the need to address *Shigella* as an urgent antimicrobial threat, in line with the WHO priority pathogen list 2024 and highlights the need to develop innovative solutions to slow transmission in sexual transmission networks where heavy antimicrobial use drives the emergence of extensively drug-resistant strains.

## Data Availability

All data produced in the present work are contained in the manuscript

## Author biography

Hannah Charles is a Principal Epidemiologist at the UK Health Security Agency. Her work focuses on the real-time and enhanced surveillance of sexually transmissible infections, including outbreaks and incidents of *Shigella*.

---

Supplementary Table 1: List of strains sequenced including NCBI Biosample accessions and their equivalent Illumina and Nanopore SRA accessions.

**Supplementary Table 2:** Characteristics of cases within t10.1814 cluster compared to the t10.377 cluster, 2016 to 2023, England.

|  |  | t10.1814<br>N=124<br>n (%) | t10.377<br>N=720<br>n (%) | P value |
| --- | --- | --- | --- | --- |
| Gender | Male | 113 (91) | 684 (95) | 0.07 |
|  | Female | 11 (9) | 32 (4) | 0.02 |
|  | Unknown | 0 (0) | 4 (0.6) | 0.39 |
| Age | Median [IQR] | 35 years [30-43] | 36 years [29-45] |  |
|  | Adult | 123 (99) | 716 (99) |  |
|  | Child | 1 (1) | 4 (1) |  |
| Travel | Yes | 20 (16) | 56 (8) | 0.004 |
|  | No/Unknown | 104 (84) | 664 (92) | 0.004 |
| Travel destination<br>(of those reporting travel) | Belgium | 1 (5) | 1 (2) | 0.05 |
|  | Brazil | 1 (5) | 0 (0) |  |
|  | Europe (country unknown) | 1 (5) | 6 (11) | 0.05 |
|  | Germany | 1 (5) | 4 (7) | 0.41 |
|  | France | 0 (0) | 2 (4) |  |
|  | Greece | 0 (0) | 1 (2) |  |
|  | Hungary | 0 (0) | 2 (4) |  |
|  | Italy | 0 (0) | 2 (4) |  |
|  | India | 1 (5) | 1 (2) | 0.05 |
|  | Netherlands | 1 (5) | 0 (0) |  |
|  | North America | 1 (5) | 0 (0) |  |
|  | Spain | 8 (40) | 24 (43) | 0.53 |
|  | South America | 0 (0) | 1 (2) |  |
|  | Turkey | 0 (0) | 1 (2) |  |
|  | Thailand | 1 (5) | 2 (4) | 0.61 |
|  | United states | 1 (5) | 2 (4) | 0.61 |
|  | Africa | 0 (0) | 1 (2) |  |
|  | Unknown | 3 (15) | 6 (11) | 0.20 |
| Presumptive MSM | Yes | 93 (75) | 626 (87) | 0.0005 |
|  | No | 31 (25) | 94 (13) | 0.0005 |
| Region | East Midlands | 4 (3) | 11 (2) | 0.48 |
|  | East of England | 2 (2) | 29 (4) | 0.28 |
|  | London | 50 (40) | 381 (53) | 0.008 |
|  | North East | 12 (10) | 9 (1) | <0.0001 |
|  | North West | 25 (20) | 90 (13) | 0.04 |
|  | South East | 16 (13) | 98 (14) | 0.77 |
|  | South West | 6 (5) | 27 (4) | 0.61 |
|  | West Midlands | 4 (3) | 26 (4) | 0.59 |
|  | Yorkshire & Humber | 5 (4) | 47 (7) | 0.21 |
|  | Unknown | 0 (0) | 2 (0.3) |  |

